# Analyzing How Changes in the Health Status of Healthcare Workers Affects Epidemic Outcomes

**DOI:** 10.1101/2020.09.21.20197574

**Authors:** I. Phadke, A. McKee, J.M. Conway, K. Shea

**Affiliations:** Department of Mathematics, The Pennsylvania State University, University Park, PA 16802; Department of Biology, The Pennsylvania State University, University Park, PA 16802; Harrisburg University of Science and Technology, Harrisburg, PA

**Keywords:** quality of care, epidemic model, mathematical modeling, healthcare workers

## Abstract

During a disease outbreak, healthcare workers (HCWs) are essential to treat infected individuals. However, these HCWs are themselves susceptible to contracting the disease. As more HCWs get infected, fewer are available to provide care for others, and the overall quality of care available to infected individuals declines. This depletion of HCWs may contribute to the epidemic’s severity. To examine this issue, we explicitly model declining quality of care in four differential equation-based SIR-type models with vaccination. We assume that vaccination, recovery, and survival rates are affected by quality of care delivered. We show that explicitly modelling healthcare workers and accounting for declining quality of care significantly alters model-predicted disease outcomes, specifically case counts and mortality. Models neglecting the decline of quality of care resulting from infection of HCWs may significantly under-estimate cases and mortality. These models may be useful to inform health policy that may differ for healthcare workers and the general population. Models accounting for declining quality of care may therefore improve the management interventions considered to mitigate the effects of a future outbreak.

## Introduction

In 2014, an Ebola epidemic ravaged western Africa. Over the course of this epidemic, healthcare access and quality became increasingly limited^1^. According to the World Health Organization (WHO), the weight of the Ebola burden caused the already limited healthcare systems of the affected countries to collapse^1,2^ and many people were not able to receive the medical aid they needed^1,3^. Specifically, quality of care provided to patients declined dramatically as an unprecedented number of healthcare workers got infected^1,4^. For example, in the Kenema Hospital in Sierra Leone, deaths among healthcare workers, already in short supply, pushed the hospital to the verge of collapse, as the Ebola epidemic progressed^5^. Clinic closures, personal protective equipment shortages and overcrowding of hospitals in the midst of the epidemic resulted from a progressively limited health force^1^. The survival rates for individuals without healthcare access were dramatically lower than they were for individuals with healthcare access^3^.

Similar tragic outcomes have been seen during the current COVID-19 pandemic. Each year dozens of potentially lethal outbreaks affect populations around the world. Annual epidemics of seasonal influenza kill thousands of people^6^, and emergent strains of pandemic influenza (e.g. the 2009 H1N1 pandemic) may have even higher mortality burden^7^. Zika emerged in the Americas starting in Brazil in 2015 and was quickly declared a public health emergency by the WHO^8^. Dozens of other infectious diseases have recently caused lethal outbreaks, including cholera, Nipah virus, Rift Valley Fever, Yellow Fever, and measles^9^. As the primary source of care, healthcare workers are essential to providing care during such outbreaks. However, their exposure means they may also become victims of the outbreak, due to their frequent contact with infected individuals. Infection hinders their ability to provide care^10^, either because policy forbids them to work, or because they are too sick to work effectively, or because they die. Access to healthcare is already a bottleneck for public health in many parts of the world^11^. As healthcare workers become infected during an outbreak, healthcare access perforce becomes limited as the system is overwhelmed, which causes the quality of care that can be offered to the average individual to decline.

Reduced quality of care may have several effects on epidemic outcomes. The absolute number of cases and disease-induced deaths could be impacted by a declining quality of care function, as healthcare workers are essential for providing vaccinations to prevent disease spread and the necessary care to reduce cases and deaths. Treating patients may reduce the duration during which the individual is infectious, and may reduce the probability that the patient dies from infection. At the population level, reduced healthcare thus likely translates into fewer administered doses of vaccine (if one exists), more transmission resulting from a longer infectious period, and increased case fatality ratios. Thus, not only does a decline in quality of care affect individual outcomes, it poses problems for controlling the epidemic at a population scale, likely increasing the number of cases, the number of deaths, and the severity and duration of the outbreak.

Conventional epidemic models fail to take into consideration potentially dynamic epidemic rates, and thus neglect the potential impact of declining quality of care^12^. As a result, such models may underestimate epidemic burdens, especially in circumstances where health system collapse is possible. The decline in quality of care over the course of a general epidemic is difficult to characterize empirically and is likely context-specific. Thus, to maintain generality, we model the quality of care function with a sigmoidal function in this paper, as this function can incorporate both linear and exponential changes to the quality of care as the proportion of uninfected individuals or uninfected healthcare workers change. We examine the effect of the specific parameterization of the quality of care function on case counts and mortality, in terms of both a loss impact parameter, which quantifies how much quality of care declines if a single healthcare worker becomes infected, and a system redundancy parameters, which quantifies what proportion of healthcare workers can be lost before quality of care sees serious decline. We compare outcomes produced by four models to assess the differences in predictions yielded by explicit consideration of healthcare as a dynamic resource, with healthcare workers either modeled explicitly or within the general population. We show that the inclusion of the quality of care may significantly impact predicted epidemic outcomes, and can therefore inform management.

## Methods

We aim to investigate the direct impact of the loss of healthcare workers (HCWs) on outbreak outcomes. To this end, we developed four SIR-type^13^ models to highlight the impact that quality of care delivered over the course of an epidemic may have on outcomes. In our full model (Fig. 1A), we distinguish health care workers from the general population, since HCWs may have higher infection rates, associated with more frequent contact with infected individuals^14^, and may also be priority targets for interventions such as vaccines^3^. We further account for the quality of care delivered by HCWs through an outbreak as a function of the uninfected fraction of the initial HCW population. In our alternate models, we relax these assumptions. For alternate model I (Fig. 1B), we assume that HCWs are a fraction of the general population rather than a distinct population, and quality of care is a function of total uninfected fraction of the population. For alternate models II and III, we neglect quality of care, but keep HCWs as a distinct population as in the main model (alternate model II, Fig. 1C) or as a fraction of the total population (alternate model III, Fig. 1D). Note that the alternate models II and III are nested within the full model and alternate model I, respectively. Thus, there are four model variants: with or without HCWs modeled explicitly, and with or without quality of care dependent on HCW status. For the two model variants in which quality of care can change, we further explore four different ways in which quality of care might decline. In the following, we outline and discuss specifically the full model and how we model quality of care as a function of healthcare worker population. Details of the remaining, alternate models are provided in the Supplementary Material.

**Figure 1:**
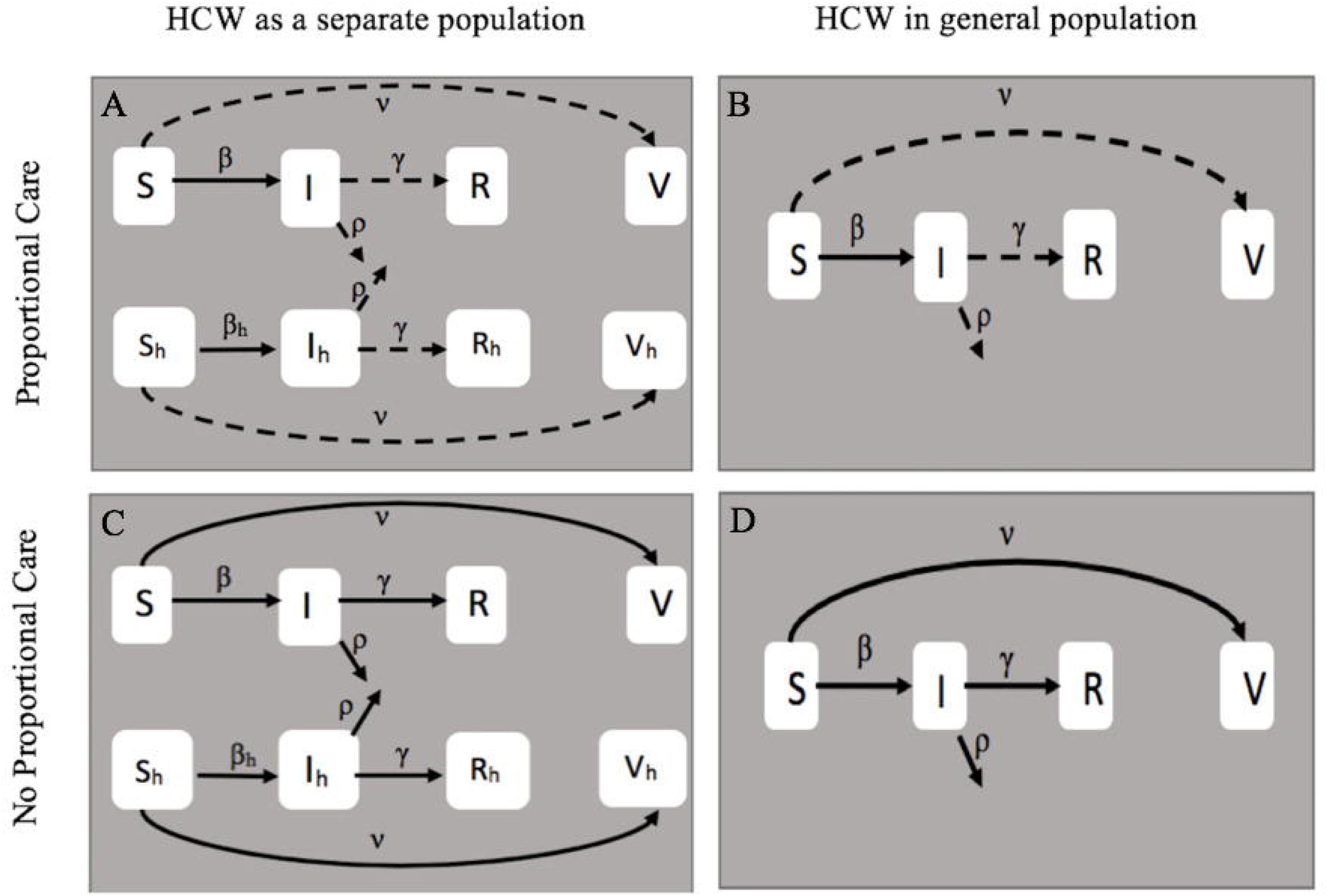
Model schematics. We visually highlight the differences between the 4 models being considered, formed by including or excluding quality of care delivered by HCWs, taking HCWs as members of the general population or a distinct population. β (Beta) is the rate of transmission, γ (gamma) is the recovery rate, ρ (survival parameter) is the probability of survival, and v (vaccination rate) is the rate of a fully effective vaccine. A: Full model, where the healthcare workers, denoted with subscript h, are a separate group from the rest of the population. Dashed lines indicate rates affected by a proportional quality of care function. B: Alternate model I, where healthcare workers are part of the general population and the dashed lines indicate rates affected by a proportional quality of care function. C: Alternate model II, where the healthcare workers are a separate group, denoted with subscript h, from the rest of the population and there is no effect of HCW loss on quality of care. D: Alternate model III where healthcare workers are part of the general population and there is no effect of HCW loss on quality of care.

### Model Assumptions

For all models, susceptible individuals, *S*, are infected at rate *β*. Infected individuals, *I*, may die with probability *ρ* or recover with probability 1-*ρ*, at recovery rate γ. We assume that once infected individuals have recovered, *R*, they cannot be re-infected. Further, we assume that vaccines also grant lasting immunity, so the susceptible individuals, vaccinated at rate *v*, will remain in the vaccinated class, *V*, at least for the duration of the outbreak. Finally, we assume that non-health care workers are vaccinated at a fraction *r* of the rate at which health-care workers are vaccinated. While many other parameters may also vary during the course of an outbreak, we keep most parameters fixed so that we can isolate the focal aspects of interest, i.e., the effects of quality of care by health care workers. Nevertheless, for completeness, we include sensitivity analyses (see the Supplementary Material). Baseline parameter values are given in Table 1.

**Table 1:** Baseline Parameters and Initial Conditions main model and alternate models I-III.

| Symbol | Parameter | Baseline value [range] | Meaning |
| --- | --- | --- | --- |
| $v$ | Vaccination rate | 0.02 weeks<sup>-1</sup> | Rate of vaccination |
| $\beta$ | Transmission rate | 1 weeks<sup>-1</sup> | Rate of transmission |
| $\beta_h$ | Transmission rate for HCW | 1 weeks<sup>-1</sup> | Rate of transmission |
| $\gamma$ | Recovery rate | 0.5 weeks<sup>-1</sup> | Rate of recovery |
| $\rho$ | Probability of Dying | 0.01 | Minimum probability of dying once infected (see text) <sup>18,19</sup> |
| $r$ | Vaccine preference | 0.5 [0,1] | Proportionate rate at which the general population is vaccinated compared to HCWs. |
| $Q(P(t))$ | Quality of Care Function | [0,1] | Quality of Care function - HCW ratio |
| $k$ | Loss impact parameter | 4.5 [0-5] | Affects how steep sigmoid function is |
| $m$ | **Redundancy parameter** | **.9 [0-1]** | Affects midpoint of sigmoid function |
| $N(0)$ | Population | 1000 | Starting total population |
| $S(0)$ | Susceptible Gen Pop | 979 | Initial number of susceptible general group |
| $I(0)$ | Infected Gen Pop | 1 | Initial number of infected general group |
| $R(0)$ | Recovered Gen Pop | 0 | Initial number of recovered general group |
| $V(0)$ | Vaccinated Gen Pop | 0 | Initial number of vaccinated general group |
| $S_h(0)$ | Susceptible Med Pop | 20 | Initial number of susceptible HCWs (see text) |
| $I_h(0)$ | Infected Med Pop | 0 | Initial number of infected HCWs |
| $R_h(0)$ | Recovered Med Pop | 0 | Initial number of recovered HCWs |
| $V_h(0)$ | Vaccinated Med Pop | 0 | Initial number of vaccinated HCWs |

### Full model

In our full model (Fig 1A), we extend a standard SIR-type model with a vaccinated class. Further, since healthcare workers (HCWs) are often subject to different dynamics, e.g. more frequent exposure to infected individuals or higher vaccination rates, we model HCWs as a separate population. Finally, we modulate vaccination, recovery, and death rates by a quality of care function, *Q*(*p*(*t*)), of the uninfected fraction, *p*(*t*), in the HCW population. We will compare predictions with alternate models I-III (Figs. 1B-1D) wherein we relax these assumptions, with alternate model III as the simplest, SIR-type model with vaccination (see Supplementary Material for details), which will permit us to explore the different dynamics resulting from the inclusion of quality of care. Differential equations describing dynamics for our full model (Fig. 1A) are given below, with all parameters defined in Table 1:

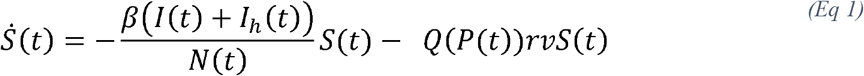

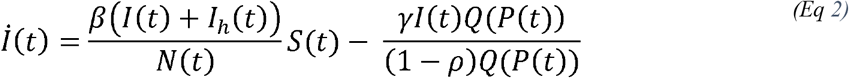

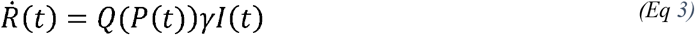

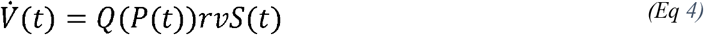

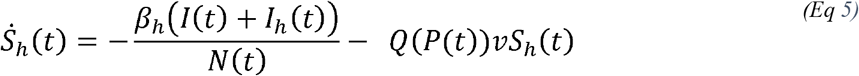

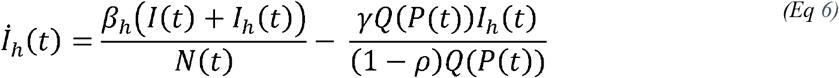

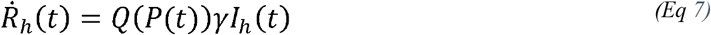

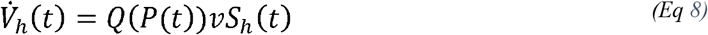

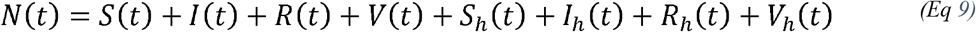

where *N* (*t*) is the total population at time t.

The full model describes a system that is overwhelmed only when healthcare workers decline. Three alternate models (Fig. 1), that include or exclude explicit quality of care functions and health care worker compartments are presented in the Supplementary Material.

### Quality of Care Function

The WHO defines of quality of care as “the extent to which health care services provided to individuals and patient populations improve desired health outcomes.” We model the quality of care, *Q* (*P*(*t*)), as a function of the proportion of HCWs, (*p*(*t*)), that are uninfected and able to provide care. Thus, for our main model, *P*(*t*)is given by the proportion of uninfected HCWs, 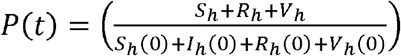. We choose *Q(P(*t*)*)to be a sigmoidal function, 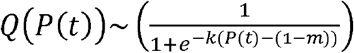, and construe the two shape parameters associated with *Q*(*P*(*t*)),*k* and *m*, as describing loss impact (*k*) and the health system redundancy (*m*). We normalize so that *Q*(*P*(*t*)) ranges from 0 to 1, which allows us to showcase healthcare systems working at levels from 0% to 100%.

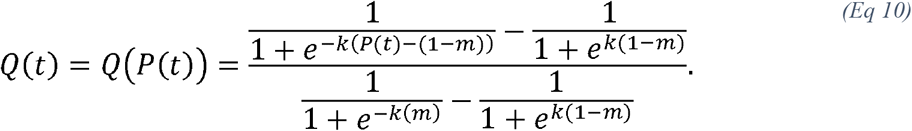

When all HCWs are infected (*p*(*t*)), the healthcare system can provide no care (Q(P(*t*))=0) and is said to be operating at 0%, while if no HCWs are infected (*P*(*t*)=1), the quality of care that the health system can provide is its best (*Q*(*P*(*t*))=1) and is operating at its full capacity of 100%. We choose a sigmoidal function because of the range of dynamics it captures via the shape parameters, m and k. Sample quality of care functions, for a few parameterizations, are provided in the Supplementary Material, with a discussion of the impact of normalization.

We relate “loss impact parameter” (*k*) to each healthcare worker’s impact on the health force system. When this parameter is smaller, a steady decline is evident in the healthcare system for each healthcare worker that is infected, while declines with HCW infected proportion are more extreme when the loss impact parameter is larger (Figs. S1 and S2). The “health system redundancy parameter” (*m*) accounts for the minimum health system provision, which is the minimum fraction of healthcare workers needed to provide adequate care. A higher health system redundancy parameter represents a healthcare system that is safeguarded compared to a lower redundancy parameter, as it is able to withstand the loss of more healthcare workers before it cannot provide adequate care (Fig. S1). When considered simultaneously, along with the normalization of the function, the two parameters interact. In systems with low loss impact, quality of care declines linearly with the proportion of healthy HCWs, regardless of the choice of redundancy parameter. The result is that quality of care is largely similar with low loss impact, and redundancy is comparatively unimportant. However, when the loss impact parameter is high, system redundancy becomes more important, and a high redundancy parameter (large *m*) overall leads a stronger quality of care provided by the health force compared to systems with a low redundancy parameter (small *m*) (Figs. S1 and S2). These issues are discussed further in the Supplementary Material.

Since the quality of care *Q*(*P*(*t*))directly depends on the number of HCWs that are able to provide care, it can modulate parameters describing the recovery rate and mortality risk for infected individuals, and population vaccination rate. We assume that the decline in quality of care will affect all of these parameters simultaneously.

### Parameter Assumptions and Initial Conditions

The complete set of initial conditions and parameters are provided in Table 1. We assume that we have a basic reproduction ratio, R_0_, of roughly 2, analogous to recent outbreaks of COVID-19, Ebola^15^, pandemic flu^16^, and SARS^17^. Further, we assume the baseline probability of dying from disease is 0.01, a value lower than that for Ebola, 0.7^18^ and SARS, 0.15^19^, but a conservative estimate of mortality rates associated with COVID-19. We assume an initial population of 1000 individuals, and that 2% of the population are health care workers in the full model and alternate model II, a representative value chosen from data provided by the WHO^20^. We assume that the outbreak is triggered by a single infected individual, and that this first case is in the general population, i.e., not an HCW.

## Results

### Underestimated Epidemic Outcomes (Cases & Deaths)

In Figure 2, we show a comparison between model-predicted cumulative infection across ranges in the redundancy parameter (*m*) and loss impact parameter (*k*) for our different model formulations. In Fig. 2A, we contrast model-predicted cumulative infections for models with no quality of care component, i.e., alternate model II (Figs. 1C; Eqs. S6-S14) (black line in Fig. 2A) with cumulative infections for all parameterizations of *m* and *k* in our models that do account for the quality of care function from the full model (Fig. 1A; Eqs. 1-9) (grey cloud in Fig. 2A). Note that, so long as both HCWs and the general population have the same parameters and the parameter for vaccine preference is set to 1, alternate model II generates identical outcomes to alternative model III (Fig. 1D; Eqs. S15-S19). The same is true for the main model and alternate model I (Fig. 1B; Eqs. S1-S5), both of which include proportional quality of care. Models that consider dynamic quality of care (full model and alternate model I) generate different outcomes than models that neglect it (alternate models II and III), regardless of parameters. Notably, the projected range of total cases increases when declining quality of care is included, leading to an increase of up to 15% in total infections (grey cloud in Fig. 2A) for the baseline parameterization (Table 1) and in the range of consideration for our loss impact (*k*) and redundancy (*m*) parameters.

**Figure 2:**
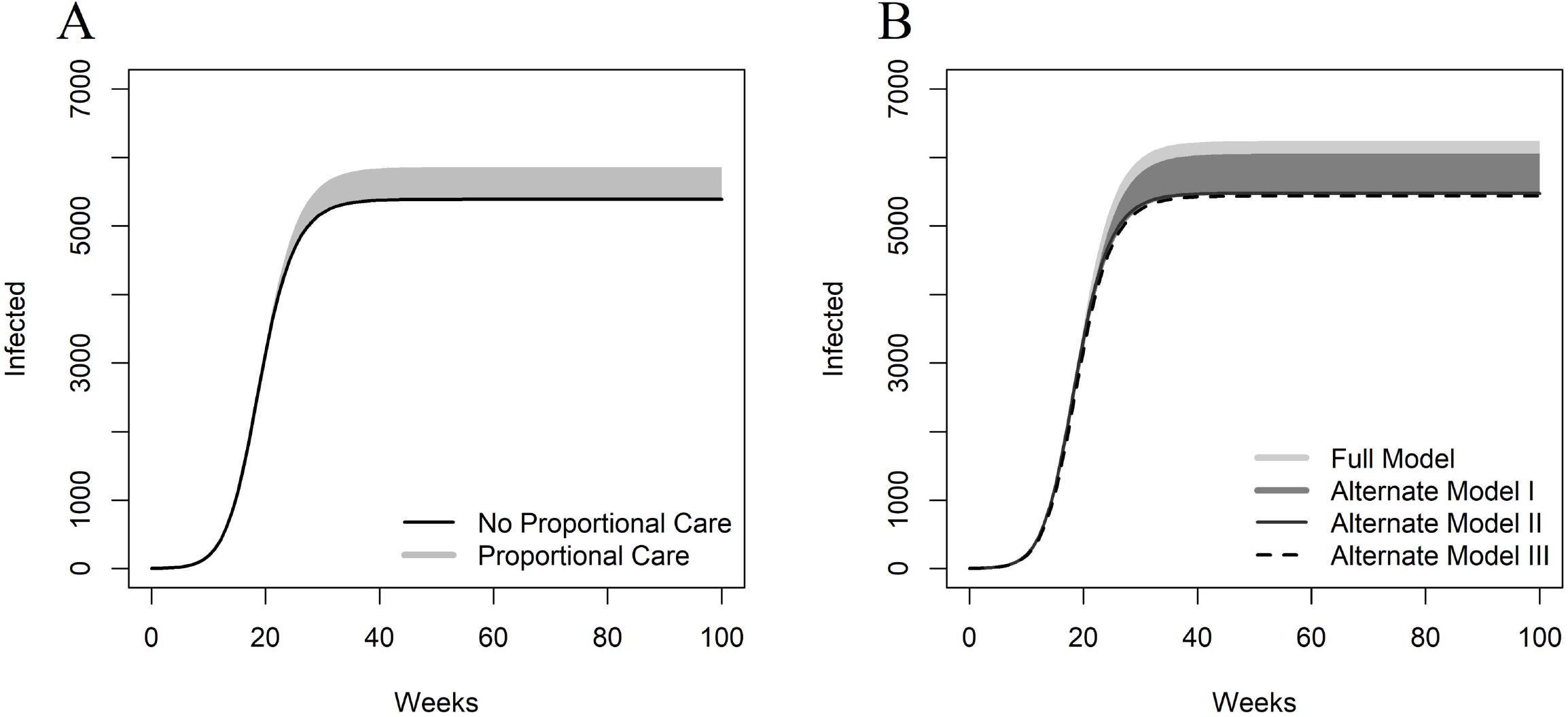
Model-predicted cumulative infections over time for different model choices. A: The grey cloud shows the range of cumulative infections that result for parameter combinations of the loss impact parameter, k=[0,5], and the redundancy parameter, m=[0,1], when a proportional quality of care function is included (Full model). The black line portrays the cumulative infections resulting from a model neglecting quality of care (Alternate model II). Parameters are the baseline parameters outlined in the table. B: The cloud shows the range of cumulative infections that result from parameter combinations of shape parameters when a proportional quality of care function is included. In this simulation, the transmission rate has been increased by 50% for healthcare workers only. This impacts the simulations for the full model and alternate model I, as HCWs are a separate population compared to the general population. All other parameters for both groups are the same.

However, HCW’s infection dynamics may differ from those of the general population. For example, they may be at greater risk of infection due to increased contact with infected individuals. In Fig. 2B, the light grey cloud (overlapped by the grey cloud), we present cumulative infections for all combinations of *k* and *m* of the quality of care function considered from the full model, while assuming infection rates are 50% higher for healthcare workers compared to the general population, achieved by assuming that the infection rate, *β*_*n*_, is 50% greater for HCWs than for the general population. We compare this to alternate model I (grey cloud in Fig. 2B), where there is a proportional quality of care based on the uninfected population, but with no differences between HCW and general population parameters, as HCW are considered to be part of the general population. A higher infection rate, associated with increased exposure to infected individuals, may explain cases in which an unprecedented number of healthcare workers have been infected^1^. Thus, the choice of quality of care function, driven by the uninfected population or uninfected HCW saturation, results in a higher number of cumulative infections when healthcare workers have a higher infection rate.

We conclude that the inclusion of quality of care has a substantial impact on the resulting epidemic outcomes (Fig. 2A).

### Impacts of Redundancy and Loss Impact Parameters on Case Counts and Deaths

Figure 3 shows the final epidemic size (Fig. 3A), mortality (Fig. 3B), and case fatality ratio (CFR) (Fig. 3C), predicted using the full model (Eqs. 1-9), assuming identical transmission rates between HCWs and the general population, *β* = *β*_*n*_ (see Table 1 for parameters), as a function of the loss impact and redundancy parameters in the quality of care function (Eq. 10).

**Figure 3:**
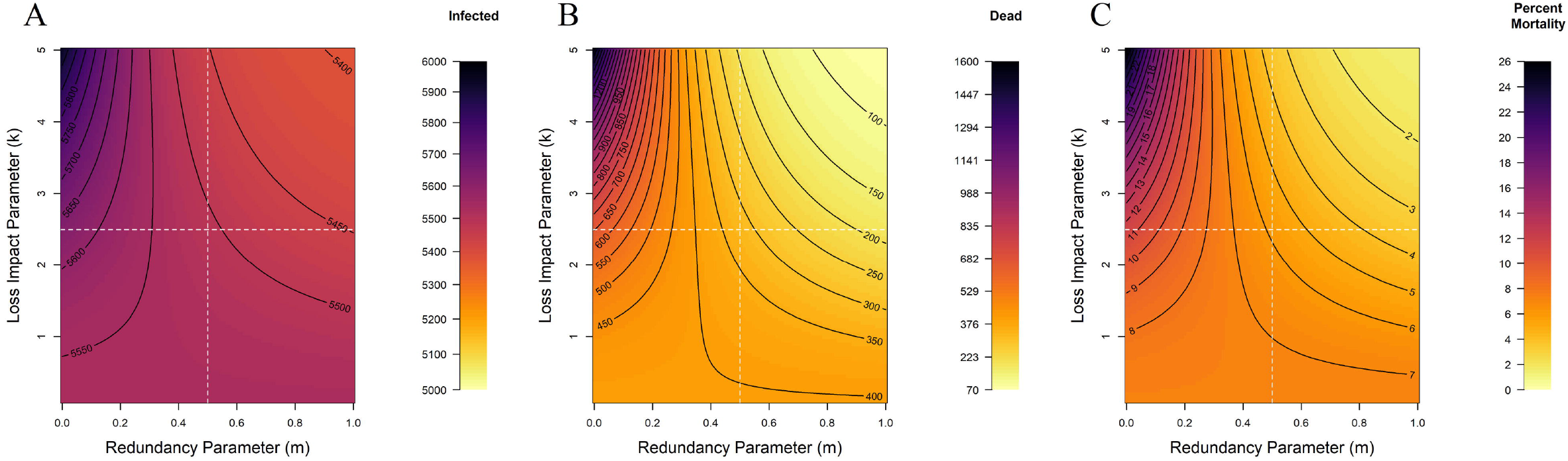
Epidemic burden resulting from each combination of the loss impact parameter k, and the redundancy parameter, m, in the quality of care function. Each panel is split into 4 quadrants representing choices of the quality of care function for the full model with the baseline parameters: A: Epidemic size is shown for each combination of the loss impact parameter and redundancy parameter in the quality of care function. B: Mortality is shown for each combination of the loss impact parameter and redundancy parameter in the quality of care function. C: Case Fatality Ratio (CFR) is shown for each combination of the loss impact parameter and redundancy parameter in the quality of care function.

Each quadrant of the contour panels in Fig. 3 relates to one of four sample quality of care functions (Fig. S1) describing: (i) low loss impact/high redundancy (modeled with low k and high m, respectively, lower right quadrant); (ii) low loss impact/low redundancy (modeled with low k and low m, respectively, lower left quadrant); (iii) high loss impact/low redundancy (modeled with high *k* and low *m*, respectively, upper left quadrant); and (iv) high loss impact/high redundancy (modeled with high *k* and low *m*, respectively, upper right quadrant). From Figure S1, we can rank the strength of a health care system based on the quality of care functions described above, from most resilient (iv) through (i) then (ii) to most fragile (iii). Thus, we investigate the impact that quality of care parameters *k* and *m*, modeling loss impact and redundancy, respectively, have on epidemic burdens. High loss impact paired with a low redundancy (upper left corner in Fig. 3A,B,C) causes the most severe epidemic outcomes with respect to case counts (Fig. 3A) and mortality (Fig. 3B), as this represents a fragile health system that has a rapid decrease in quality of care, *Q(P(t*)), as few healthcare workers are infected (*Q (P(t*)) the dotted black line in Fig. S1). Conversely, the mildest epidemic predictions result from the combination of high system redundancy and high loss impact (upper right corner in Fig. 3A, B, C), as this represents a strong health system, due to the normalization used to obtain the quality of care function (see Fig. S2). This latter prediction arises is because there is a minimal decrease in quality of care until nearly all healthcare workers are infected, when there is a rapid decrease in quality of care. As that many healthcare workers are rarely infected, this means quality of care is rarely significantly reduced.

High loss impact describes a large decline in performance of the healthcare system as an additional healthcare worker is infected, after some threshold determined by the redundancy parameter. Thus, high loss impact paired with low redundancy (modeled by small *m*) means that the healthcare system is not well safeguarded, so a few infections amongst HCWs will cause the system to collapse. If this combination is representative of a current system, that system can mitigate epidemic burdens by either reducing the loss impact parameter (perhaps through cross-training of HCWs) or increasing redundancy in the system. It is important to note that the strongest systems have high loss impact and high redundancy; if both parameters are controllable, our modeling predicts that a focus on improving redundancy would offer the most effective improvements in quality of care. For the mildest predicted epidemic outcomes, in cases where loss impact is high, our modeling predicts that a higher level of redundancy would yield a healthcare system robust enough to withstand a large number of infections prior to rapid collapse only once a very high proportion of HCWs are infected.

### Parameters Impacted by Infected HCWs

As an epidemic progresses, inclusion of the dynamic quality of care function clearly leads to drastic differences compared to static quality of care, i.e., taking *Q(P(*t*)*) ≡1 (Fig. 4A). When decline in the quality of care is considered in the model, the number of cumulative deaths increase by 1716%, the number of vaccinations administered decreases by 10.8%, and the number of individuals that recover decreases by 8.9%. The quality of care used in Fig. 4A is one that is representative of a weak health care system with a high loss impact and low redundancy (Fig. S1, dotted black line). Figure 4B shows the impact inclusion of dynamic quality of care has on cumulative deaths for the four general quality of care functions described above (Fig. S1) and compares it to the resulting epidemic burdens when quality of care is static. While the predicted increase in cumulative deaths that result from modeling quality of care in Fig. 4B are not all as severe as seen in Fig. 4A, the inclusion does further showcase the impact its consideration has on epidemic burdens.

**Figure 4:**
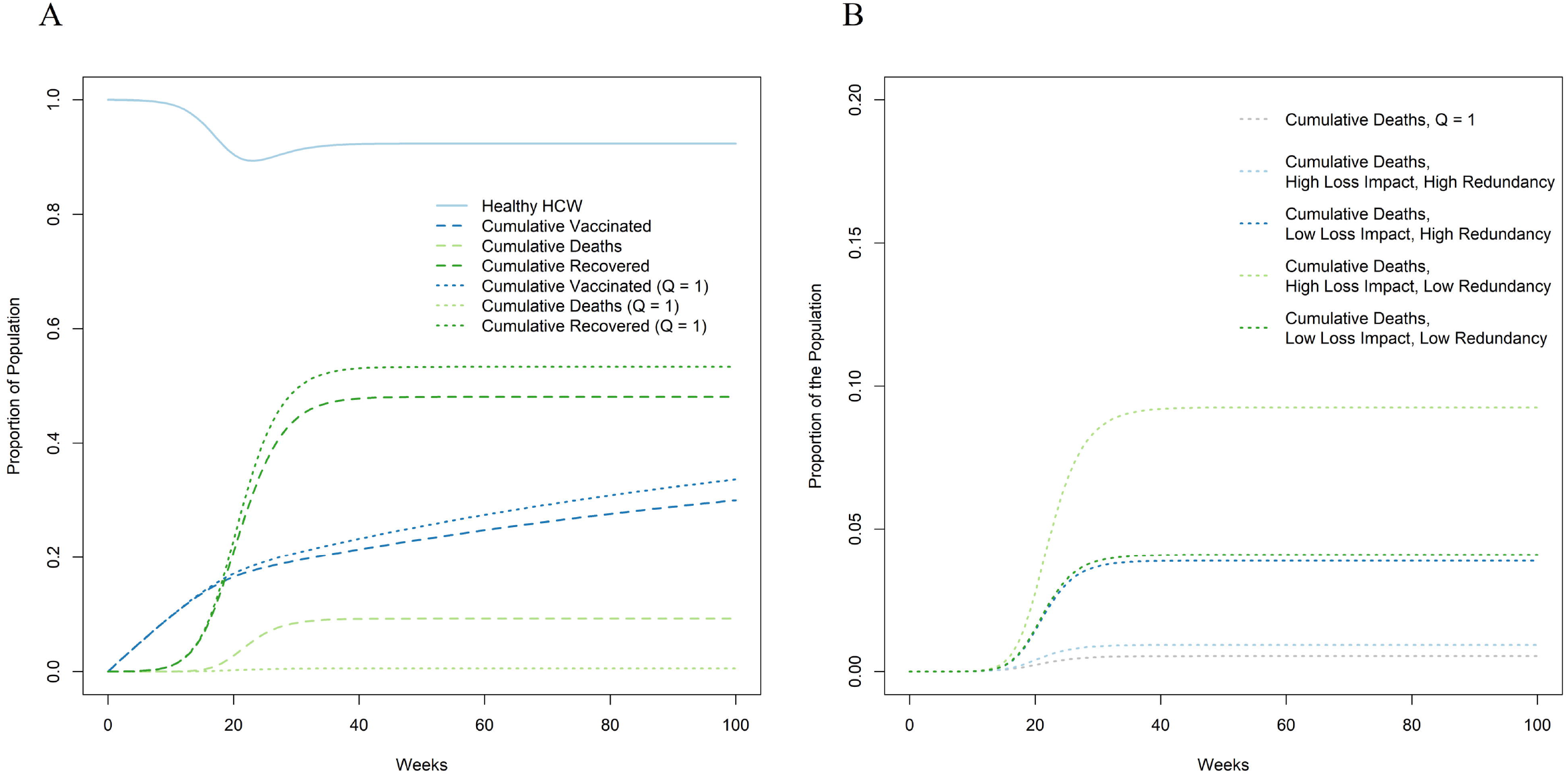
Impact of dynamic quality of care on cumulative infections and mortality predictions, due to changes in the infected HCW proportion. A: Comparison of resultant cumulative infections when comparing rates affected by the proportional quality of care function: vaccination rate, recovery rate and likelihood of death between a dynamic quality of care model and when the quality of care function is set to be maximal (*Q(P (*t*)*) ≡ 1), equivalent to models that do neglect it. Lines for Cumulative Vaccinated, Recovered and Deaths are representative of the impact the infected HCWs have on those parameters for simulation with the full model with baseline parameters. The model considered uses a quality of care function representing a weak healthcare system, i.e. high loss impact (*k* = 4.5) and low redundancy parameter (*m* = 0.1). Lines representing rates followed by Q=1 are representative of the assumption quality of care is 1, represented by simulation with alternate model II with baseline parameters. B: Comparison between sample quality of care functions and their resulting cumulative deaths per simulation. Cumulative deaths resulting from the simulation for the four sample sigmoidal quality of care functions from Figure S1 (see Supplementary Material) are shown in addition to the number of deaths resulting from having the quality of care *Q(P (t*)) ≡ 1, equivalent to models that do neglect it. This subfigure highlights the range of differences in cumulative deaths resulting from the shape of the quality of care function considered.

## Discussion

Conventional epidemic models generally do not consider quality of care that changes as a disease outbreak proceeds, yet loss of healthcare workers and other impacts on quality of care could potentially radically affect disease dynamics, as it may have in the 2014 Ebola outbreak^1,21^, and as seen in the current COVID-19 pandemic. In this paper, we demonstrate that evolving quality of care impacts disease dynamics, and that inclusion of declining quality of care negatively impacts epidemic burdens.

If quality of care is assumed to be static in a predictive model, that model may dramatically underestimate epidemic burdens. This underestimate is also dynamic; a model assuming static quality of care that fits well early in an outbreak will diverge in predictions later in the outbreak. The magnitude of the impact that declining quality of care has on disease dynamics depends strongly on the way in which quality of care declines in response to the proportion of HCWs affected; a wide range of relatively mild to severe epidemic impacts may arise. Explicit consideration of healthcare worker dynamics (in contrast to assuming HCWs are well-mixed with the general population) also generally leads to more severe outcomes, suggesting that modeling them separately may be necessary. Failing to include either of these aspects in epidemic models may cause one to underestimate, in some cases severely, epidemic burdens, especially in contexts where HCWs are a limiting factor and likely to experience different dynamics than the rest of the population.

Knowing the empirical response of the healthcare system to loss of HCWs during the course of a serious outbreak would be ideal, but such trajectories are likely highly dependent on both context and pathogen. We avoid this issue here by assuming that quality of care declines according to different, but plausible, functional forms. We specifically consider the loss impact (each healthcare worker’s impact on the health system) and the level of built-in redundancy in the health system (the minimum number of health care workers required to provide adequate care); both are important, but redundancy dominates outcomes, as seen in Supplementary Material Fig. S2. Nevertheless, it is possible to learn about quality of care decline for a particular situation. For example, data on HCW infection and death rates in the initial part of the COVID-19 pandemic in China, or in Europe might reasonably have been extrapolated to the situation in other parts of the world with similar health systems even before COVID-19 reached those other nations.

Models that explicitly incorporate dynamic quality of care may aid decision-making to control an epidemic. By anticipating the degree to which declining care quality might impact disease dynamics, control methods may be tailored to mitigate a healthcare system’s redundancy and/or factors affecting loss impact, depending on the existing structure of the healthcare system. For example, the redundancy component of our quality of care function did dominate outcomes and therefore is a likely initial target for improvements. This could be useful for management; for example, the level of effort put in to recruiting/training healthcare volunteers could be increased, which in turn would cause the system’s redundancy to improve. In the current COVID-19 pandemic, movement of medical personnel from less-affected locations, recruitment of recently retired HCWs, and accelerated graduation of final year medical students have all been used to sustain the health care system in pandemic hotspots. It also may be beneficial to manage healthcare workers differently from the rest of the population to attempt to slow the decline in quality of care. Arguments are already made to prioritize healthcare workers for vaccination and/or other medical interventions as they are the primary source of care in epidemics^22^ and often are faced with higher rates of infections compared to the general population^23^. Careful exploration of other dynamic quality of care parameterizations may also lead to the identification of alternative management interventions. For example, information about quality of care decline may inform outside interventions, such as how many and when HCWs from NGOs are usefully to be deployed to assist in times of crisis, as seen in health care worker shortage during the Kenema Hospital collapse^5^ (e.g. in terms of when system collapse is anticipated). This model is capable of distinguishing between these strategies, and thus is able to estimate the efficacy of a wider range of potential strategies.

As with any model, there are caveats. We have used a relatively simple base model to assess the general insights that arise from a first consideration of plausible declines in quality of care in epidemic settings; more complex model structures, and parameterization for specific diseases are warranted in future work. We also assume that other factors that might vary during an epidemic are constant to isolate the effect of quality of care decline. For example, future work should explore: that HCW infection rates may decline (if infection prevention and control are improved once a pathogen is recognized) or increase (as Personal Protective Equipment (PPE) becomes scarce, as in COVID-19 in some settings); the effects of different case fatality ratios, and of HCW recovery, in more complex models; the impact of prioritized vaccination of HCWs; next generation calculations of R_0_; different contact structures in the HCW and general population; and, the context of outbreaks in health care systems in high income vs low/middle income countries with inherent differences in health care system structures. We used a sigmoidal function to describe quality of care; in the future, we could consider other functional forms bounded between 0 and 1, for example a Hill function^24^, or a piecewise linear function. It is important to note that quality of care is likely context specific, due to the global heterogeneity in healthcare access; results may thus depend on local circumstances. Furthermore, the way in which the most complex models differentiate HCWs from the general population is deserving of further exploration; we did not fully consider how HCWs differ in infection risk and care received. As mentioned earlier, empirical data would be immensely informative, but difficult to obtain in advance. Nevertheless, with sufficient information it will be possible to generate an empirically supported quality of care function for a specific disease outbreak, and use it to examine likely dynamics for future outbreaks of the same disease in similar health care settings, and avoid the problems inherent in a just-in-time response. It might also be possible to address multiple subclasses of HCWs (e.g. physicians, nurses, janitorial staff, midwives, volunteers) with different quality of care responses and different transmission dynamics based on their roles^25^. This would further allow us to explore various prioritization methods for healthcare workers to minimize impacts on quality of care provided^26^.

The quality of care function might also be improved by including information on healthcare workers’ behavioral responses to the status of an epidemic. As an epidemic progresses further, the personal risk of working increases for healthcare workers. Behavioral avoidance of risk by HCWs, especially volunteer HCWs, may accelerate system collapse in extreme situations^21^. Applying game theoretic approaches may usefully address these tensions between personal and population-level benefits and costs/risks as HCW risk preferences may vary over the course of an epidemic^27,28^. Due to the severity and nature of the HIV outbreak in the 1980s, there were cases where healthcare workers felt overwhelmed, and in some cases avoided practicing, due to fears and burdens of the disease^29^. Accounting for behavioral or related tendencies which may impact healthcare worker participation in the work force is essential to consider their impacts to quality of care provided.

Our present work focuses on the healthcare system as it pertains to the management of a generic disease, but clearly there are ramifications for the quality of care provided for other public health issues. As healthcare declines, levels of care for other patients, affected by other issues, will also suffer^30^. For example, health seeking-behavior for malaria was impacted by concerns about Ebola^31,32^. During the COVID-19 pandemic, “non-essential” surgeries and procedures are being canceled in anticipation of hospital surges or out of fear of infection; with significant indirect ramifications for the health of those patients, and ironically leading to HCW layoffs in some cases (which damages health system redundancy). In this case, the implications of failing to consider a declining quality of care function would be compounded and models that account for it could be used to address a potential major gap in standard epidemiological modelling approaches.

In conclusion, we here demonstrate that consideration of the potential for declining quality of care affects epidemic outcomes, in some cases quite significantly. Addressing such declines in quality of care explicitly will allow epidemiologists to anticipate, plan for and mitigate changes in epidemic dynamics over the course of an epidemic. While much work remains to be done, addressing these critical issues offers significant potential public health benefits. Given the crisis posed by the current COVID-19 pandemic, the declines in quality of care arising from HCW illness and death^33^, and the lack of appropriate PPE and medical equipment such as ventilators that also reduces quality of care in many health care settings, such careful consideration is warranted.

## Data Availability

n/a

## Acknowledgements

We would like to thank the Millennium Scholars Program at Penn State and Joe Keller, Emily Howerton, Hidetoshi Inamine, Jack Seifarth, Trine Ready, Bella Ruud, Elyse Johnson, and other Shea Lab members for their thoughtful comments and guidance throughout the course of this project.

We also acknowledge the Applied Math REU 2016 at Penn State and Dean Mary Beth Williams for the funding they provided to IP. Additionally, this work was partially supported by funding from the National Science Foundation RAPIDD Grant DMS-1514704, NIH EEID Award 1 R01 GM105247-01 and NSF COVID-19 RAPID award DEB-2028301 (KS) and by National Science Foundation Grant No. DMS-1714654 and NIH grant 1 R21 AI143443-01A1 (JMC).

